# Automatic Population of the Case Report Forms for an International Multifactorial Adaptive Platform Trial Amid the COVID-19 Pandemic

**DOI:** 10.1101/2023.09.19.23295797

**Authors:** Andrew J. King, Lisa Higgins, Carly Au, Salim Malakouti, Edvin Music, Kyle Kalchthaler, Gilles Clermont, William Garrard, David T. Huang, Bryan J. McVerry, Christopher W. Seymour, Kelsey Linstrum, Amanda McNamara, Cameron Green, India Loar, Tracey Roberts, Oscar Marroquin, Derek C. Angus, Christopher M. Horvat

## Abstract

**Objectives:** To automatically populate the case report forms (CRFs) for an international, pragmatic, multifactorial, response-adaptive, Bayesian COVID-19 platform trial.

**Methods:** The locations of focus included 27 hospitals and 2 large electronic health record (EHR) instances (1 Cerner Millennium and 1 Epic) that are part of the same health system in the United States. This paper describes our efforts to use EHR data to automatically populate four of the trial’s forms: baseline, daily, discharge, and response-adaptive randomization.

**Results:** Between April 2020 and May 2022, 417 patients from the UPMC health system were enrolled in the trial. A MySQL-based extract, transform, and load pipeline automatically populated 499 of 526 CRF variables. The populated forms were statistically and manually reviewed and then reported to the trial’s international data coordinating center.

**Conclusions:** We accomplished automatic population of CRFs in a large platform trial and made recommendations for improving this process for future trials.

## Introduction

Randomized-controlled trials (RCTs) are the cornerstone of evidence-based medicine yet are remarkably resource-intensive^1,2^. Numerous efforts are ongoing to reduce the per-result cost of RCTs, including improving patient recruitment^3^, developing trial designs that can simultaneously test multiple conditions or treatments^4^, applying Bayesian statistics to detect superiority/inferiority more quickly^5^, or creating reusable trial infrastructure^6,7^. One area ripe for reducing RCT costs is automating the traditionally manual task of completing case report forms (CRF).

CRFs are paper or electronic forms in which each trial participant’s clinical characteristics and outcomes are recorded^8^. These forms are sent from the site where an enrolled patient is participating to a data coordinating center responsible for calculating trial outcomes. Usually, CRFs are completed by clinical research coordinators who collect the data required to populate a trial’s CRFs. In contemporary clinical trials, most of the required data are manually extracted from the participants’ electronic health records (EHRs).

Because EHRs are often the primary data source that clinical research coordinators use when populating CRFs, it is theoretically possible to automate the data extraction process and automatically populate many variables within a trial’s CRFs. However, due to the complexity of healthcare, the intricacies of EHR documentation practices, and the mismatch between the periodically documented variables present in an EHR and the authoritative (and sometimes multifaceted) variables required by CRFs, automating this task (outside of retrospective proof of concept studies^9,10^) has mainly been unattainable.

### Objective

Given the urgency of the COVID-19 pandemic, there was a desire to enroll as many patients as possible. We quickly realized that the availability of trained personnel to coordinate trial administration and collect trial outcomes would be a rate-limiting factor. To increase our enrollment potential, we aimed to automatically populate the trial’s CRFs from the two different EHR systems used at 27 hospitals participating in this platform trial. A second objective was to build expertise and learning health system infrastructure to be redeployed in future trials.

## Methods

### Locations, data sources, and trial design

At the start of the pandemic in April 2020, UPMC (a quaternary care medical center headquartered in Pittsburgh, Pennsylvania) began enrolling patients for an international platform trial^11^. The primary EHR system at most of UPMC’s hospitals was Cerner Millennium; however, six hospitals used Epic. To support the trial, a secure research environment was created to store and process identifiable patient data. The data for patients enrolled at Cerner hospitals were queried and duplicated in their raw form. The data for patients enrolled at Epic hospitals were queried and duplicated from an Epic Clarity database.

During the planning phase, trial data requirements were broadly categorized into structured and unstructured (**Figure 1**). Unstructured elements included 1) the majority of eligibility questions because potential patient enrollees are not guaranteed to have had sufficient, preceding interactions with the health system to support scraping comprehensive eligibility data from the electronic record; 2) protocol deviations, adverse events, and serious adverse events as these instances were deemed to warrant research team adjudication of events via review, in part, of clinical notes; 3) an adjudication log, which kept track of updates made to correct for noise and clarify EHR data; 4) specific patient characteristics, such as days of symptoms prior to illness or whether the patient’s occupation was as a healthcare worker, which were required for CRF reporting but not available as standalone fields in the health system’s EHRs; and 5) long-term outcomes which were collected by research coordinators via a phone survey. Structured data elements included patient characteristics such as age and other demographic information readily retrievable from the EHR relational databases, as well as elements of care such as vital signs, laboratory values, organ support therapies and settings, and medication administrations available as tabular data. Unstructured elements were collected as structured data using separate web application forms for screening, enrollment, recording of long-term outcomes, and documentation of protocol deviations and adverse events, which were made accessible to research coordinators. Additional unstructured elements, such as symptoms and comorbidities, were captured as structured data using a COVID-19 ‘intake form’ made available to clinicians for completion in Cerner and Epic EHRs. The data change log was maintained within the trial reporting database by the study data team.

**Figure 1.**
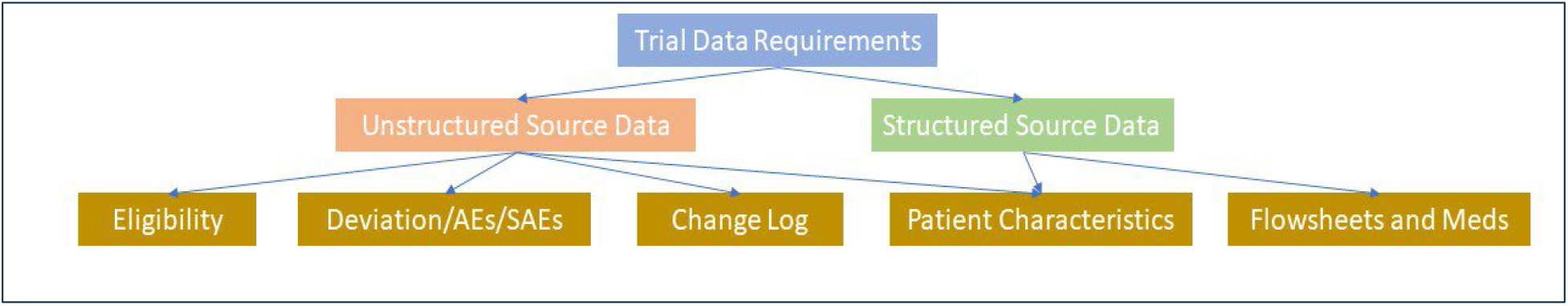
Trial data reporting requirements were categorized broadly as structured and unstructured data. Unstructured data elements were captured with electronic forms integrated into either research coordinator or clinician workflows. Structured data were collected from the EHRs relational databases. AE, adverse event; SAE, serious adverse event.

The randomized, embedded, multifactorial, adaptive platform trial for community-acquired pneumonia and COVID-19 (REMAP CAP: COVID-19) is an innovative clinical trial that provides a backbone trial structure upon which multiple intervention ‘domains’ can be simultaneously tested^4^. Domains are categories of treatments and are comprised of interventions that belong to that category. For example, low-molecular-weight heparin was a treatment in the anticoagulation domain^12^ and hydrocortisone was a treatment in the corticosteroid domain^13^. Individual patients can be assigned to a combination of treatments across multiple domains. The trial relies on Bayesian statistics to periodically update randomization weights as the trial progresses and data accrue, such that enrolled patients are increasingly assigned to treatment arms most likely to offer benefits.

REMAP CAP was instantiated to study community-acquired pneumonia in critically ill adult patients. The trial designers had the foresight to craft a pandemic appendix to the core protocol should a pandemic emerge while the trial was being conducted. In April 2020, REMAP CAP activated this appendix, and REMAP CAP: COVID-19 was launched in the US to study interventions for treating patients with COVID-19, with a primary outcome of organ support-free days^4^. Additionally, trial entry criteria were expanded to include non-critically ill adults hospitalized outside intensive care units (ICUs).

### Case report forms

This paper focuses on our efforts to use EHR data to automatically populate four forms: baseline characteristics, daily outcomes, discharge outcomes, and response-adaptive randomization characteristics and outcomes. These four forms represent 41% of all data collected for each patient. (Data reported for each enrolled patient included 13 CRFs covering 1,289 variables.) We focus on these forms for automation because they were the most reliant on structured clinical data collected during the hospital stay but also required careful curation and transformation to meet the reporting requirements of the trial. While not the focus of this manuscript, additional views were created to support the manual completion of other forms, including those for adverse events, serious adverse events, and protocol deviation. We did not automate the adverse event and protocol deviation forms because they often required a review of unstructured clinical notes, which we chose not to include. Instead, we prioritized what we could accomplish with structured data.

### Development environment

The secure environment was only accessible from within the firewall of the health system and using computers managed by the health system. The environment had MySQL installed as its database management system. The research team did not have administrative permissions to install additional software, so the decision was made to develop the entire pipeline using MySQL queries. The result of the MySQL queries was a series of four tables corresponding to the four different CRFs of focus with embedded logic checks highlighting areas requiring adjudication (for example, data indicating a patient was simultaneously receiving multiple types of respiratory support). A clinician informaticist (author CMH) continuously adjudicated data from the MySQL pipeline through chart review to ensure accuracy. The adjudicated data were used to inform and validate code updates. This was necessary to ensure both code and resultant data validity while accounting for the rapidly evolving trial, with domains and interventions opening and closing as new ideas to treat COVID-19 emerged and as tested treatments proved efficacious or not. These tables were exported to CSV files where a set of quality assurance scripts written in the R statistical computing language were run on the local computer before the CSV files were securely transferred to the international data coordinating center using Globus (research cyberinfrastructure).

### Development strategy

The emergent and dynamic nature of the pandemic necessitated a code development strategy that could keep pace with a complex and rapidly evolving clinical trial, to achieve the mission of collecting data from disparate EHRs with elaborate underlying databases. An iterative, rapid application development framework was followed, with cycles determined by data deadlines to support response adaptive analysis updates, safety reporting requirements, and trial domain analyses. This approach allowed an expedited planning period necessary to meet external demands while incorporating robust feedback via data validation into each development cycle. Early cycles involved adjudication of all EHR-extracted data elements with chart review to ensure the code was functioning as intended; later cycles involved chart review validation of any data elements collected via updated code and all data related to the primary trial outcome of organ support free days at trial day 28.

## Results

Between April 4, 2020 and May 6, 2022, 417 patients were enrolled in the trial at UPMC. The MySQL-based extract, transform, and load pipeline was successful in automatically populating 95% of the clinical variables present on four CRFs, including 188 of 192 variables on the form for baseline characteristics, 113 of 113 variables on the form for daily outcomes, 109 of 132 variables on the form for discharge outcomes, and 89 of 89 variables on the form for response-adaptive randomization characteristics and outcomes. Of the automatically populated variables, 75% (374/499) were sourced directly from the structured source data, while 25% (125/499) were sourced from the unstructured source data. Of the variables that were not automatically populated, five were related to the location of lung infiltrates as determined by chest imaging, and the rest were related to adverse events, including major bleeding, myocardial infarction, deep vein thrombosis, and pulmonary emboli.

Pipeline development underwent three major versions, each becoming more comprehensive and accurate based on feedback from clinician adjudication by comparison of the completed CRFs with electronic record source documentation. The first version of the pipeline was organized around a patient’s ICU admission, as the trial’s primary outcomes focused on days of organ support. Collected data occasionally required annotated correction to account for patients with multiple ICU admissions during a single hospitalization or transfers between hospitals within the health system, the latter generated a new set of account and encounter identifiers for an enrolled patient.

The second version of the pipeline was organized around a patient’s hospitalization. All the data for a patient’s hospital stay (including external transfers) were harmonized, and derived tables were created for tracking a patient’s unit location (i.e., emergency department, ICU, stepdown, ward) and receipt of organ support included in the trial primary outcome (i.e., high flow nasal cannula, non-invasive mechanical ventilation, invasive mechanical ventilation, extracorporeal membrane oxygenation, and vasopressor medications). This effort required close coordination with our health system’s ‘ICU Service Center’ operations leadership group, which helped the study team closely track the fluctuating status of units that were temporarily converted to either ICUs or combined ICU-ward ‘COVID-19 units.’ Separate queries were created for the Cerner Millennium database and the Epic Clarity database, as well as for the two different patient enrollment states as defined by the trial, which consisted of a ‘moderate’ state for patients not requiring organ support at the time of enrollment and a ‘severe’ state for patients receiving organ support at the time of enrollment. The queries for these four groups of patients (Cerner-moderate, Cerner-severe, Epic-moderate, Epic-severe) included over one thousand lines of code each, all with significant maintenance requirements.

The third version was a rewrite of version two to optimize the architecture, reduce maintenance overhead, and facilitate code reuse. This version was constructed with an improved data model informed by lessons learned during the initial implementation of the pipeline and as familiarity developed with the trial. First, the source tables (from both Cerner and Epic) would flow into the same curated tables with harmonized patient identifiers, locations/unit stays, standardized event names, values, units, value sets, and time zones. From these tables, a single set of scripts transforms the data into the necessary derived tables regardless of source EHR or patient enrollment state. Finally, information from the curated and derived tables was combined into tables representing four different CRFs (**Figures 2 and 3**).

**Figure 2.**
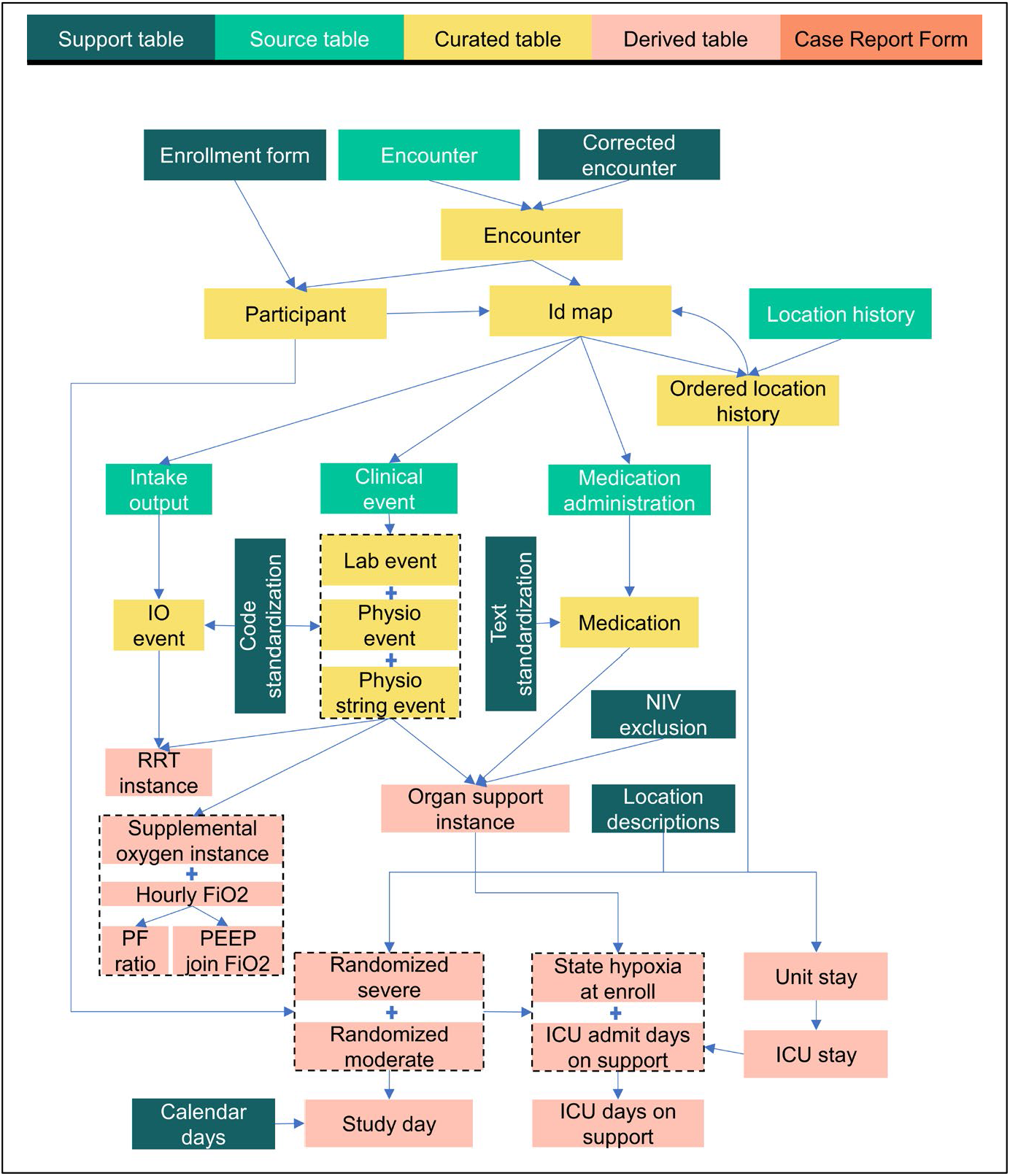
Information flow for automatic population of a clinical trial’s case report forms: part 1. This figure shows how source tables containing raw EHR data and manually curated support tables are combined to create a series of curated tables that harmonize and standardize participant data. These tables are then combined to create a series of derived tables. Note, temporary tables, such as those needed to support temporal joins of different rows of the same table, are not shown.

**Figure 3.**
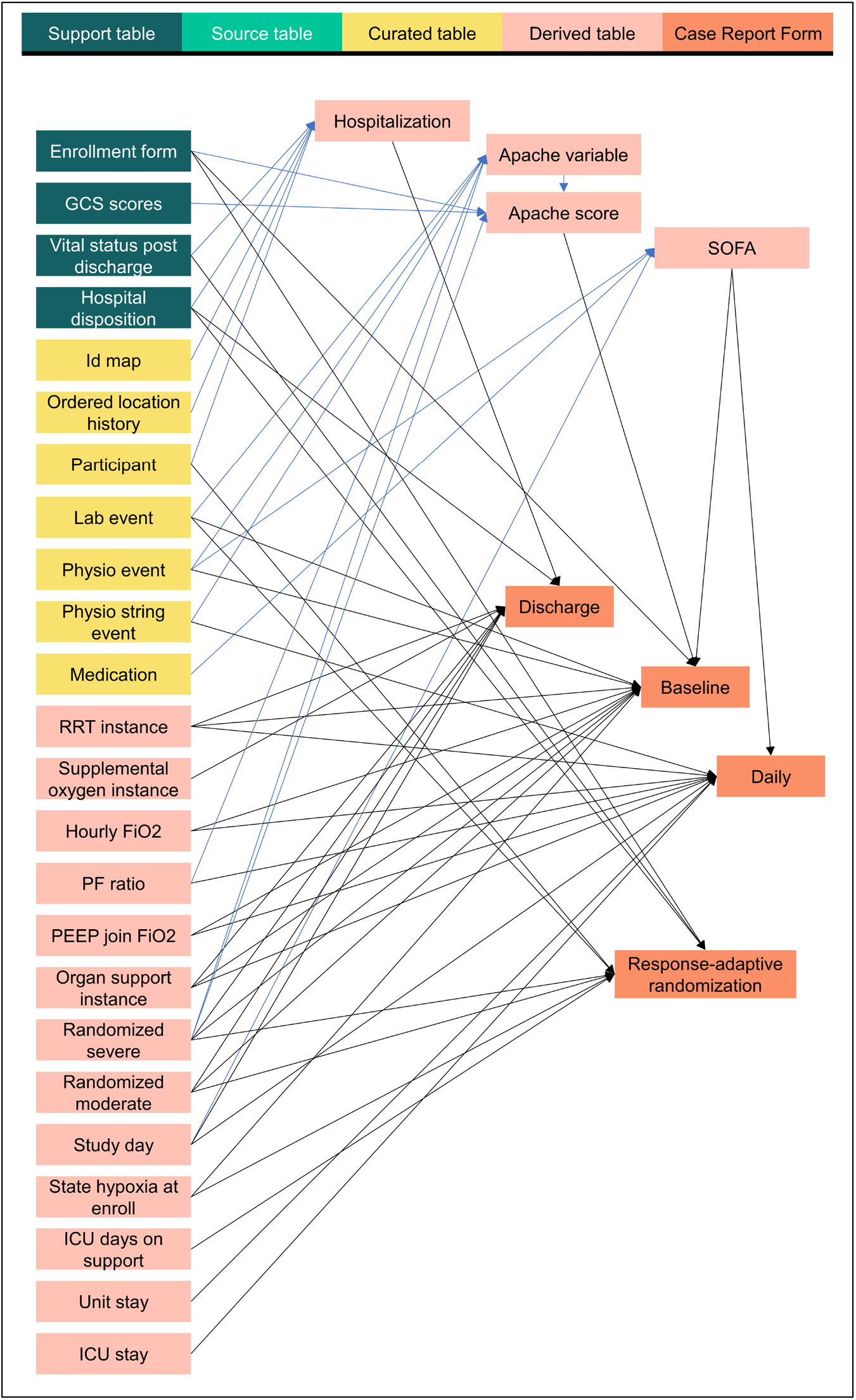
Information flow for automatic population of a clinical trial’s case report forms: part 2. This figure shows how support tables, curated tables, and derived tables are joined to populate the four tables corresponding to the CRFs for baseline characteristics, daily outcomes, discharge outcomes, and response-adaptive randomization characteristics and outcomes.

A sample of the challenges we faced, our solutions (from pipeline version three), and recommendations for future projects are provided in **Table 1**.

**Table 1.**
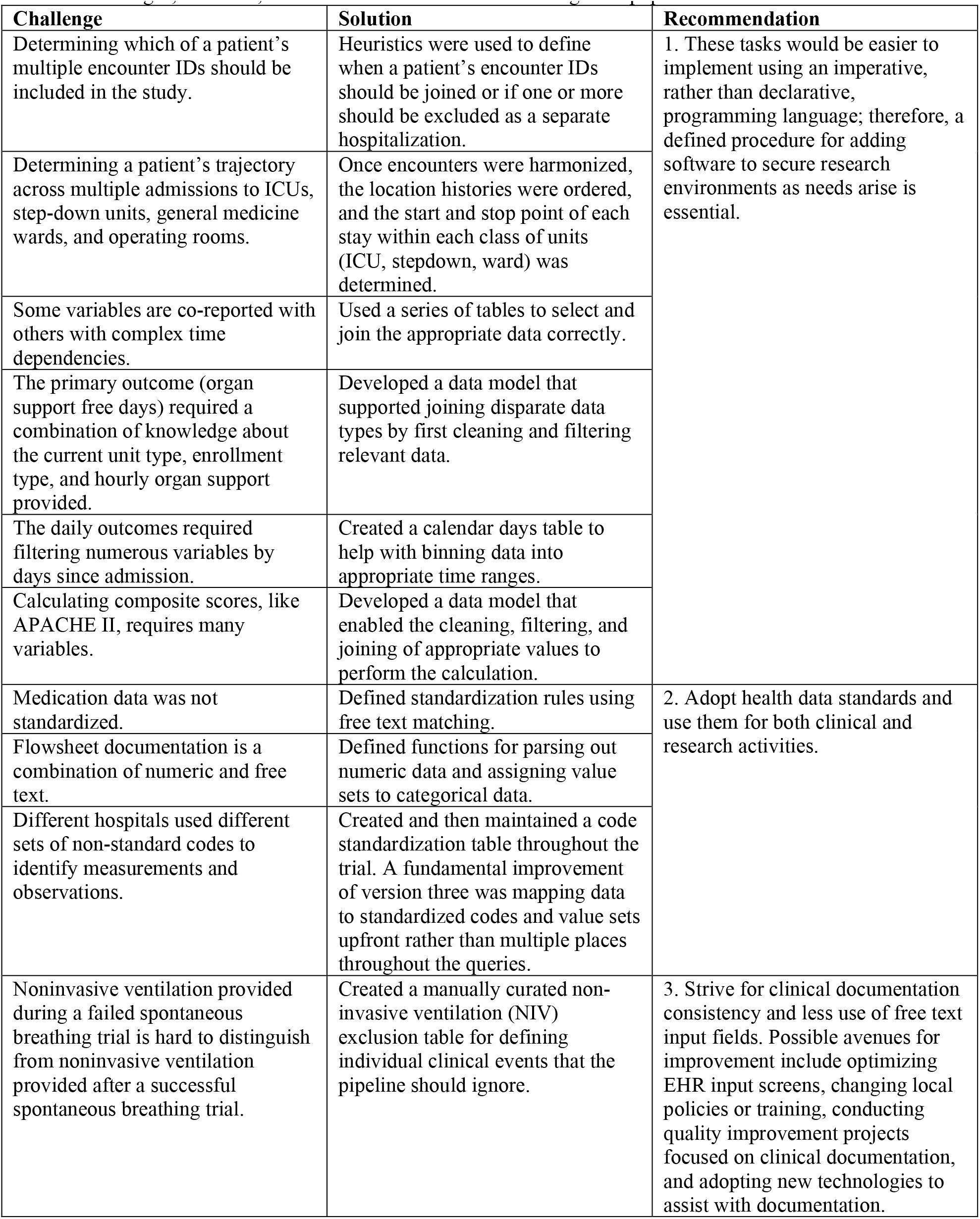
Challenges, solutions, and recommendations for automating CRF population from EHR data.

| **Challenge** | **Solution** | **Recommendation** |
| --- | --- | --- |
| Determining which of a patient's multiple encounter IDs should be included in the study. | Heuristics were used to define when a patient's encounter IDs should be joined or if one or more should be excluded as a separate hospitalization. | 1. These tasks would be easier to implement using an imperative, rather than declarative, programming language; therefore, a defined procedure for adding software to secure research environments as needs arise is essential. |
| Determining a patient's trajectory across multiple admissions to ICUs, step-down units, general medicine wards, and operating rooms. | Once encounters were harmonized, the location histories were ordered, and the start and stop point of each stay within each class of units (ICU, stepdown, ward) was determined. |  |
| Some variables are co-reported with others with complex time dependencies. | Used a series of tables to select and join the appropriate data correctly. |  |
| The primary outcome (organ support free days) required a combination of knowledge about the current unit type, enrollment type, and hourly organ support provided. | Developed a data model that supported joining disparate data types by first cleaning and filtering relevant data. |  |
| The daily outcomes required filtering numerous variables by days since admission. | Created a calendar days table to help with binning data into appropriate time ranges. |  |
| Calculating composite scores, like APACHE II, requires many variables. | Developed a data model that enabled the cleaning, filtering, and joining of appropriate values to perform the calculation. |  |
| Medication data was not standardized. | Defined standardization rules using free text matching. | 2. Adopt health data standards and use them for both clinical and research activities. |
| Flowsheet documentation is a combination of numeric and free text. | Defined functions for parsing out numeric data and assigning value sets to categorical data. |  |
| Different hospitals used different sets of non-standard codes to identify measurements and observations. | Created and then maintained a code standardization table throughout the trial. A fundamental improvement of version three was mapping data to standardized codes and value sets upfront rather than multiple places throughout the queries. |  |
| Noninvasive ventilation provided during a failed spontaneous breathing trial is hard to distinguish from noninvasive ventilation provided after a successful spontaneous breathing trial. | Created a manually curated non-invasive ventilation (NIV) exclusion table for defining individual clinical events that the pipeline should ignore. | 3. Strive for clinical documentation consistency and less use of free text input fields. Possible avenues for improvement include optimizing EHR input screens, changing local policies or training, conducting quality improvement projects focused on clinical documentation, and adopting new technologies to assist with documentation. |

## Discussion and Conclusions

Automatically populated CRFs were used during analyses that led to multiple practice-changing publications, including those providing evidence related to the COVID-19 treatments of therapeutic anticoagulation^12,14^, hydrocortisone^13^, interleukin-6 receptor antagonists^15^, convalescent plasma^16^, antiplatelet therapy^17^, angiotension converting enzyme inhibition and angiotensin receptor blockage^18^, lopinavir-ritonavir, and hydroxychloroquine^19^. In this manuscript, we presented the MySQL data model and affiliated pipeline used to report data for the patients enrolled at UPMC in the United States, a subset of the larger group of participating locations.

Reflecting on lessons learned, we have compiled desiderata for reducing ambiguity in a trial’s CRF completion guidelines (see **Table 2**) from the perspective of informaticians to facilitate automation when drawing clinical trial data from real-world data sources. These recommendations aim not to set strict requirements but to list practices that will facilitate quicker automation of CRF pipelines. Following these items will save time by reducing the communication needed between trial sites and data coordinating centers.

**Table 2.** Desiderata for reducing ambiguity in a trial’s CRF completion guidelines.

|  |  |
| --- | --- |
| **a.** | **Create an EHR-CRF variable mapping section and index.** CRF completion guidelines can be hundreds of pages long. All the fields and the EHR variables required for populating those fields should be listed in a single section of the document, which can serve as an index pointing to the pages of the document where each variable is referenced. |
| **b.** | **All fields and variables should be given a unique identifier.** Creating such identifiers will reduce ambiguity when discussing individual variables. It can create consistency in how variable names are encoded in processing scripts across trial sites, the statistical analysis committee, and the data coordinating centers. |
| **c.** | **Variable definitions and priority.** When defining a field such as a baseline serum sodium measurement, the laboratory tests that are acceptable to use should be listed along with the priority of using them. For example, choosing between whole blood and serum sodium values collected closely together in time. |
| **d.** | **Include standardized codes and related value sets, and rely on interoperable code whenever able.** When defining variables, value sets of acceptable standardized codes should be included. For example, the LOINC codes 2951-2, 42570-2, and 77139-4 all refer to Sodium measurements and may be included in the acceptable set of values. As interoperable data storage formats, such as the Observational Medical Outcomes Partnership common data model, become more widespread at academic institutions affiliated with health systems, and as EHR vendors work to comply with the United States Core Data for Interoperability requirements, sharing interoperable queries and adopting federated approaches to data sharing promises to streamline work in this space. |
| **e.** | **Define acceptable units and allowable values.** Each variable should include preferred and acceptable units and the range of acceptable values. Discrete fields should include value sets of acceptable responses. |
| **f.** | **Provide explicit time bounds.** The EHR purview can reach far back in time for an enrolled patient. The CRF completion guidelines should specify the earliest acceptable timepoint (e.g., $\leq 24$ hours of enrollment but not before the current hospitalization), the latest acceptable timepoint (e.g., $\leq 2$ hours after enrollment), and the priority of values when multiple are available (e.g., the measurement closes to the enrollment time or the first available measurement). |
| **g.** | **Provide explicit sequence and time bounds for co-reported fields.** Interpretation of some fields, such as PaO<sub>2</sub>, FiO<sub>2</sub>, and PEEP, may require values to be reported at similar points in time. When this is the case, any sequence requirements and time bounds, or lack thereof, should be specified (e.g., with the earliest available value taking priority, the timestamps of values for FiO<sub>2</sub> and PEEP must be $>0$ and $\leq 120$ minutes after the timestamp of each reported measurement for PaO<sub>2</sub>). Such time bounds are helpful when programming variable alignment that may be intuitive to clinicians reading a chart. |
| **h.** | **Be consistent with definitions.** Adaptive trials may include separate reporting forms for updating the randomization function and calculating endpoint results. When variables on these forms overlap, they should be given precisely the same definition. If this is impossible, the variables should be clearly distinguished in the variable definition section. |
| **i.** | **Utilize a common platform for document design and dissemination.** The pandemic gave rise to virtual platforms that facilitate collaboration amongst individuals separated by wide-ranging geographies. A shared development platform allows for the ready exchange of ideas and allows trial teams to keep up to date with CRF versions and other important documents. |

Our recommendations are complementary to others working in this space, for example, making electronic CRFs more findable, accessible, interoperable, and reusable (FAIR) to support better reuse of clinical research data^20^ and auditing if planned data sharing aligns with post-publication reality^21^. A final advantage of defining a standard data model for atomic, individual patient-level data is that the code for transforming curated data into derived variables can be shared across participating sites and reused in multiple trials^22^.

### Conclusions

We described our successful effort to automatically populate CRFs from EHR data for a clinical trial amid a global pandemic. Additionally, we present desiderata for designing CRF completion guidelines to support researchers implementing auto CRF in future trials. Emerging interoperability standards hold the promise of substantially streamlining work in this area. However, widespread adoption of these standards will take time and may not meet all the requirements of a given trial. Early engagement of informaticians with deep knowledge of local EHR documentation practices and their representation in backend databases will contribute to the success of future efforts aiming to automate CRF extraction.

## Data Availability

The data underlying this article cannot be shared publicly due to institutional policies
that protect the privacy of individuals whose data were used in the study.

## Acknowledgments

Dan Rickets, Jonathan Holton, Kevin Quinn. This effort was funded in part by the UPMC Learning While Doing Program.

